# The decline of renal function aggravates arterial stiffness: a large-scale cross-sectional and longitudinal study

**DOI:** 10.1101/2024.01.01.24300697

**Authors:** Qiuping Zhao, Wei Wang, Yiming Leng, Jie Wang, Miao Rujia, Zhiheng Chen, Jiangang Wang, Jingjing Cai, Hong Yuan, Fei Li, Yao Lu

**Author notes:** **Corresponding to Yao Lu**, Health Management Center, the Third Xiangya Hospital, Central South University, 138 Tongzipo Road, Changsha, 410013, China. Wang and Zhao contributed equally to this work.

## Abstract

**Purpose:** Many studies investigated the one-single-direction relationship between arterial stiffness and chronic kidney dysfunction, particularly in patients with end-stage renal disease. The bidirectional relevance between kidney function decline and arterial stiffness in general population remains unknown. This study aimed to address the temporary relationship between arterial stiffness and renal function.

**Materials and Methods:** This large-scale observational study comprised one cross-sectional and one longitudinal population sample totalling 67,060 individuals aged over 18 years with brachial-ankle pulse wave velocity (baPWV) and estimated glomerular filtration rate (eGFR) measurements available. Associations with potential risk conditions were analysed using multiple regression analyses. Cox proportional model was used to investigate the association of arterial stiffness and incident chronic kidney disease (CKD). Cross-lagged path analysis was further conducted to analyze the temporal relationship between baPWV and eGFR.

**Results:** Multiple regression analyses showed that baPWV was inversely associated with eGFR. Compared with the lower baPWV tertile group, higher baPWV was a predictor of CKD risk, with increased HRs for three baPWV tertile groups [HR=2.17 (1.26-3.76), P for trend <0.05]. Accordingly, lower eGFR was significantly associated with higher arterial stiffness risk, even after full-adjusted [HR=1.21 (1.02-1.44), P for trend <0.05]. In the path analysis, the coefficient of the association between baseline baPWV and follow-up eGFR was lower than the effect of baseline eGFR for follow-up baPWV (−0.063 Vs. −0.077, P <0.001).

**Conclusions:** Decrease of eGFR appeared to aggravate arterial stiffness, which unravelling a new understanding of the role kidney dysfunction played in arterial stiffening.

## INTRODUCTION

Chronic kidney dysfunction is associated with dramatically increased cardiovascular mortality.^1–3^ This excess cardiovascular risk is in part attributed to an increase of traditional risk factors and may also relate to the complex metabolic and vascular changes, including arterial stiffening.^4^ Arterial stiffness, as measured by pulse wave velocity (PWV), is a strong marker of vascular aging.^5^ Recent studies showed that the measurement of arterial stiffness could improve the predictive strength of cardiovascular disease, in contrast to traditional risk factors.^6,7^ Arterial stiffness and chronic kidney dysfunction are closely linked by shared risk factors and associated increased cardiovascular mortality.

Many studies have investigated the cross-sectional associations between PWV and kidney function, with inconsistent results.^8–10^ Several studies showed the independent impact of arterial stiffness on renal function decline in patients with chronic kidney diseases, particularly in those with end-stage renal disease. The existed evidence with inconsistent results is based on people with kidney dysfunction and also lacks the bidirectional study to clarify this association.^8–10^ Understanding the temporal associations of PWV and renal function in the general population may provide an early prediction and medication of both vascular aging and chronic kidney diseases.

Here, we present both cross-sectional and longitudinal associations between arterial stiffness and renal function decline and address the temporary relationship in general adults, using a large cross-sectional study and a longitudinal study from China, which total an unprecedented 67060 individuals.^11^

## MATERIALS AND METHODS

### Study Population

The study samples were recruited from the Health Management Center of the Third Xiangya Hospital (Changsha, Hunan Province), the largest physical examination institution in central China, and also the Vascular Health Management Project research center. Hundreds of institutions came to the hospital for regular medical examinations every year, which guaranteed the high follow-up rate for this study. Patients from communities and rural areas came for vascular function examination is also a part of the study samples. Details of the study population were published in https://doi.org/10.1016/j.jacc.2019.12.039.11

In the cross-sectional study, we included 67828 participants aged over 18 years during 2012-2016. Participants without brachial-ankle pulse wave velocity (baPWV) measurements (n=51), as well as creatinine information (n=254), or with severe kidney disease (defined as eGFR<15 ml/min per 1.73 m^2^, n=23; and/or microalbuminuria >300 mg/ml, n=463) were excluded from the study, with 67060 individuals finally in the analysis. In the cohort study, we extended excluded the participants without a follow-up record of baPWV and creatinine (n=54736), resulting in the last sample size of 12324 (Figure 1). All of the participants were given informed consent. The study protocols were approved by the Medical Ethics Committee of the Third Xiangya Hospital of Central South University.

**Figure 1.**
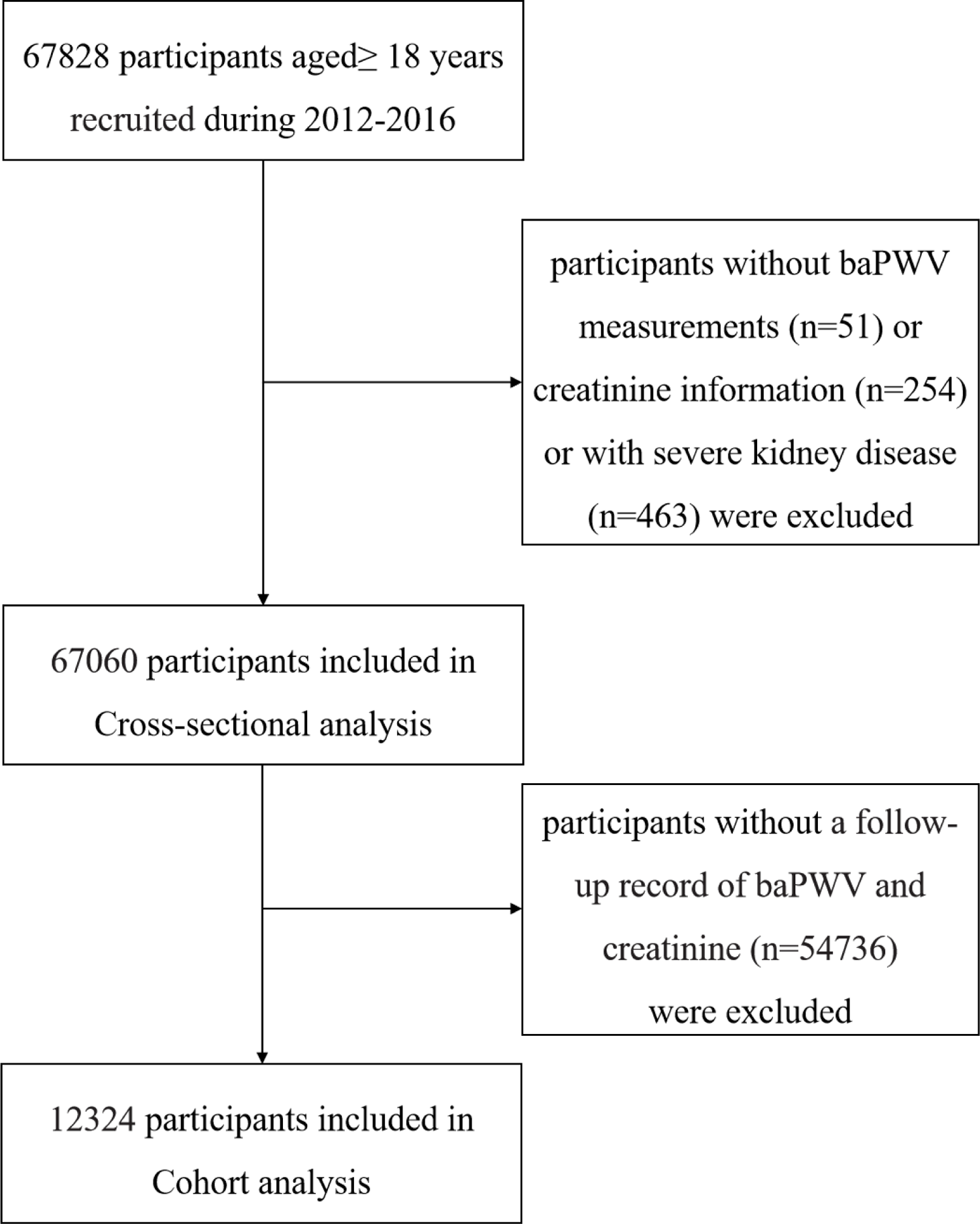
The flow chart of the observational study

### Data Collection

A standardized questionnaire had been conducted by uniform trained staff to collect the personal information of all participants (including age, sex, smoking, drinking, physical activity, and disease history of hypertension and diabetes) at each visit. Based on extensive literature, we defined these indicators as follows: Current smoking was defined as continuous cigarette consumption >1 per day in recent 6 months. Current drinking was defined as continuous alcohol consumption (liquor, beer, or wine) >2 days per week in the recent 12 months. Physical activity was defined as whether with regular mild to moderate physical activity ≥30 min at least 3 times per week. Bodyweight and height were measured wearing light clothes without shoes by an automated instrument, to the nearest 0.1 kilograms or 0.1 centimeters, respectively. Body-mass index (BMI) was calculated as weight in kilograms divided by the square of the height (kilograms per meter squared). Heart rate and blood pressure (BP) were measured repeated twice with 10 minutes rest interval, using an electronic sphygmomanometer. The average value of the two-times records was used as the final record. Mean arterial pressure (MAP) was calculated as: MAP=diastolic BP + [1/3 (systolic BP - diastolic BP)], where systolic BP and diastolic BP were in mmHg. Hypertension was defined as participants with self-reported diagnosed hypertension history and/or with a BP value of ≥140/90 mmHg. Diabetes was defined as participants with self-reported diagnosed diabetes history and/or fasting glucose ≥7.0 mmol/L.

### baPWV Measurement

The baPWV was measured by two trained physicians using a non-invasive vascular screening device (BP-203 RPE Ⅲ, Omron health medical (Dalian, China) Co., LTD). Detailed measurement procedures had been described previously.^11^ Measurements were conducted in the checkroom with a comfortable temperature between 22-25℃. Smoking and coffee drinking are prohibited before measurements. After 5-minutes of rest, participants were asked to take off their coats and socks, wearing light clothes lie on the examination couch. The electrocardiogram electrodes and heart sound sensor were placed first. Four cuffs were put on the upper arms and ankles on both sides, which connected to the plethysmographic sensor and oscillometric pressure sensor, for pressure waveforms measurements. The baPWV was measured twice for both sides. The average mean values were used as the final record.

### Laboratory measurements

Fasting venous blood samples were collected from each participant after fasting for 8-10 hours in the morning, which were immediately centrifuged at 3000×g for 10 minutes and stored at −80°C until analysis. Morning fasting midstream urine samples were collected from each participant and frozen at −40°C. Triglyceride (TG), low-density lipoprotein (LDL), high-density lipoprotein (HDL), serum creatinine, fasting blood glucose (FBG), and urinary levels of creatinine, and albumin were measured using an automatic biochemical analyzer (model 7600; Hitachi, Ltd., Tokyo, Japan). Details of these assays were described previously.^11–13^

### Assessment of Renal Function

Renal function was assessed with eGFR and microalbuminuria. eGFR was calculated using the formula adapted from the Modification of Diet in Renal Disease equation based on data from Chinese subjects with CKD. The specific formula is as follows: eGFR=175 × serum creatinine^-1.234^ × age^-0.179^ (× 0.79 for girls/women), where serum creatinine concentration is in milligrams per deciliter and age is in years. The presence of CKD was defined as eGFR < 60 ml/min per 1.73 m^2^ or elevated microalbuminuria of at least 30 mg/ml.^14^

### Statistical analysis

All statistical analyses were performed with Stata software version 16.0 (Stat Corp., College Station, Texas, USA), SPSS 22.0 (SPSS Inc., Chicago, IL, USA), and SPSS Amos 22.0 (SPSS Inc., Chicago, IL, USA). Continuous variables were used Shapiro–Wilk test indicated a non-normally distribution. The significance of differences between groups was conducted with Kruskal–Wallis test for continuous variables and the chi-square test for categorical variables. Two-side P values of <0.05 were considered significant in all analyses. Cox proportional hazards models were applied to compare the risk of incident CKD across the baPWV groups. Strengths of the effect were determined by the hazard ratios (HRs) and their 95% confidence intervals (95% CIs). Cross-lagged panel analysis was used to investigate the temporal relationship between arterial stiffness and eGFR changes.

## RESULTS

### Cross-sectional association of eGFR and baPWV

Totally 67060 participants (61662 without CKD; 5389 with CKD) were enrolled in the cross-sectional study, with a median age of 47.0 (interquartile range, IQR: 39.0-54.0). Males accounted for 61.0%. The average baPWV level in the CKD group is 1492.5 (1334.5-1709.0), while in the non-CKD group is 1138.0 (1218.0-1491.0) (P <0.05) (Supplemental Table 1).

In the cross-sectional analysis, we used multiple regression analysis to examine the effects of conventional cardiovascular risk factors on baPWV and other factors independently associated with baPWV. After stepwise adjusted for confounders, factors independently associated with baPWV were age, sex, BMI, physical activity, heart rate, systolic BP, FBG, TG, and eGFR (Table 1). eGFR was negatively associated with baPWV (β=-0.208, P<0.001, Table 1). We also explored the effects of the risk factors with eGFR as shown in Table 2. A similar significant negative association was observed between baPWV and eGFR (β=-0.003, P<0.001, Table 2).

**Table 1.** Multiple regression analyses of factors that affect arterial stiffness (baPWV)

| Variables | Model 1 | Model 2 | Model 3 |
| --- | --- | --- | --- |
| Age | 12.953* | 12.382* | 9.300* |
| Male | 54.497* | 65.573* | 28.203* |
| BMI | 6.097* | 5.737* | -6.031* |
| Current smoking |  | -8.991 | 1.917 |
| Current drinking |  | 3.044 | 4.475 |
| Physical activity |  | -16.596* | -13.7403* |
| Heart rate |  |  | 4.067* |
| MAP |  |  | -0.502 |
| SBP |  |  | 7.194* |
| FBG |  |  | 12.507* |
| TG |  |  | 7.717* |
| HDL |  |  | 6.327 |
| eGFR |  |  | -0.208* |

Data are standard regression coefficients (values) and level of significance. \* $P < 0.001$ .
Abbreviation: BMI, body mass index; eGFR, estimated glomerular filtration rate; FBG, fasting blood glucose; HDL, high-density lipoprotein; MAP, mean arterial pressure; SBP, systolic blood pressure; TG, triglyceride.

**Table 2.** Multiple regression analyses of factors that affect kidney function (eGFR)

| Variables | Model 1 | Model 2 | Model 3 |
| --- | --- | --- | --- |
| Age | -0.643* | -0.604* | -0.590* |
| Male | -20.486* | -20.335* | -18.728* |
| BMI | -0.280* | -0.373* | -0.300* |
| Current smoking |  | 0.869* | 0.945* |
| Current drinking |  | 2.348<sup>#</sup> | 1.640 |
| Physical activity |  | -0.837<sup>#</sup> | -1.372* |
| Heart rate |  |  | 0.175* |
| MAP |  |  | -0.124* |
| SBP |  |  | 0.097* |
| FBG |  |  | 1.979* |
| TG |  |  | 0.240 |
| HDL |  |  | 5.643* |
| baPWV |  |  | -0.003* |

Data are standard regression coefficients (values) and level of significance. \* P<0.001.
Abbreviation: BMI, body mass index; eGFR, estimated glomerular filtration rate;
FBG, fasting blood glucose; HDL, high-density lipoprotein; MAP, mean arterial pressure; SBP, systolic blood pressure; TG, triglyceride.

### Longitudinal association of baseline eGFR and baPWV

In the longitudinal analysis, firstly, to explore the bidirectional association between CKD and arterial stiffness. We exclude the participants with baseline diagnosed with CKD to investigate the association of baPWV and CKD risk (n=947). The prevalence of incident CKD in baPWV tertile groups from low- to high was 18.68%, 27.11%, and 54.11%, respectively (P for trend <0.001). Compared with the lower baPWV group, higher baPWV was a predictor of CKD risk, with increased HRs for three tertile groups (P for trend <0.05, Table 3). After full adjustment, the significant association of baPWV and incident CKD in the highest baPWV tertile still existed with HR of 2.17 (1.26-3.76) (Table 3). Accordingly, the lowest eGFR was significantly associated with higher arterial stiffness risk in the full-adjusted model [HR=1.21 (1.02-1.44), Model 3, Table 4], after excluded participants with baseline diagnosed with arterial stiffness to investigate the association of eGFR and arterial stiffness risk (n=5042).

**Table 3.** Associations of baPWV with incident CKD.

| Model | Tertile 1 | Tertile 2 | Tertile 3 | P for trend |
| --- | --- | --- | --- | --- |
|  | baPWV<1272.5 | 1272.5≤baPWV<1432 | baPWV≥1432 |  |
| Case, n (%) | 51 (18.68) | 74 (27.11) | 148 (54.11) | <0.001 |
| Crude | 1.00 | 1.37 (0.96-1.95) | 2.51 (1.83-3.45) | <0.001 |
| Model 1 | 1.00 | 1.16 (0.78-1.73) | 1.72 (1.17-2.54) | 0.006 |
| Model 2 | 1.00 | 1.19 (0.76-1.86) | 1.74 (1.13-2.68) | 0.012 |
| Model 3 | 1.00 | 1.53 (0.89-2.63) | 2.17 (1.26-3.76) | 0.006 |

Cox proportional model was used to analyze the association between baPWV and CKD risk. P for trend test was calculated by Spearman's correlation for continuous variables and Cochran–Armitage trend test for binary variables.
Model 1: adjusted for baseline age, gender, BMI;
Model 2: further adjusted for current smoking, current drinking, physical activity;
Model 3: further adjusted for heart rate, SBP, MAP, FBG, TG, HDL. Abbreviation:
baPWV, brachial-ankle pulse wave velocity; BMI, body mass index; FBG, fasting blood glucose; HDL, high-density lipoprotein; MAP, mean arterial pressure; SBP, systolic blood pressure; TG, triglyceride.

**Table 4.** Associations of eGFR with incident arterial stiffness.

| Model | Tertile 1 | Tertile 2 | Tertile 3 | P for trend |
| --- | --- | --- | --- | --- |
| | eGFR $\geq 124.4$ | $104.2 \leq \text{eGFR} < 124.4$ | eGFR $< 104.2$ | |
| Case,n(%) | 465 (27.79) | 563 (33.65) | 645 (38.55) | <0.001 |
| Crude | 1.00 | 0.85 (0.77-0.96) | 0.68 (0.60-0.76) | <0.001 |
| Model 1 | 1.00 | 1.20 (1.06-1.36) | 1.27 (1.18-1.58) | <0.001 |
| Model 2 | 1.00 | 1.16 (1.01-1.34) | 1.32 (1.11-1.56) | 0.001 |
| Model 3 | 1.00 | 1.11 (0.96-1.29) | 1.21 (1.02-1.44) | 0.032 |

Cox proportional model was used to analyze the association between baPWV and CKD risk. P for trend test was calculated by Spearman's correlation for continuous variables and Cochran–Armitage trend test for binary variables.
Model 1: adjusted for baseline age, gender, BMI;
Model 2: further adjusted for current smoking, current drinking, physical activity;
Model 3: further adjusted for heart rate, SBP, MAP, FBG, TG, HDL. Abbreviation:
baPWV, brachial-ankle pulse wave velocity; BMI, body mass index; FBG, fasting blood glucose; HDL, high-density lipoprotein; MAP, mean arterial pressure; SBP, systolic blood pressure; TG, triglyceride.

### Temporal relationship between eGFR changes and baPWV

Figure 2 presents a cross-lagged path analysis of arterial stiffness and renal function progression. The path coefficient from baseline eGFR to the follow-up baPWV (β1=-0.077; P<0.001) was significantly greater than the path coefficient from baseline baPWV to the follow-up eGFR (β2=-0.063; P<0.001), with P<0.001 for the difference between β1 and β2.

**Figure 2.**
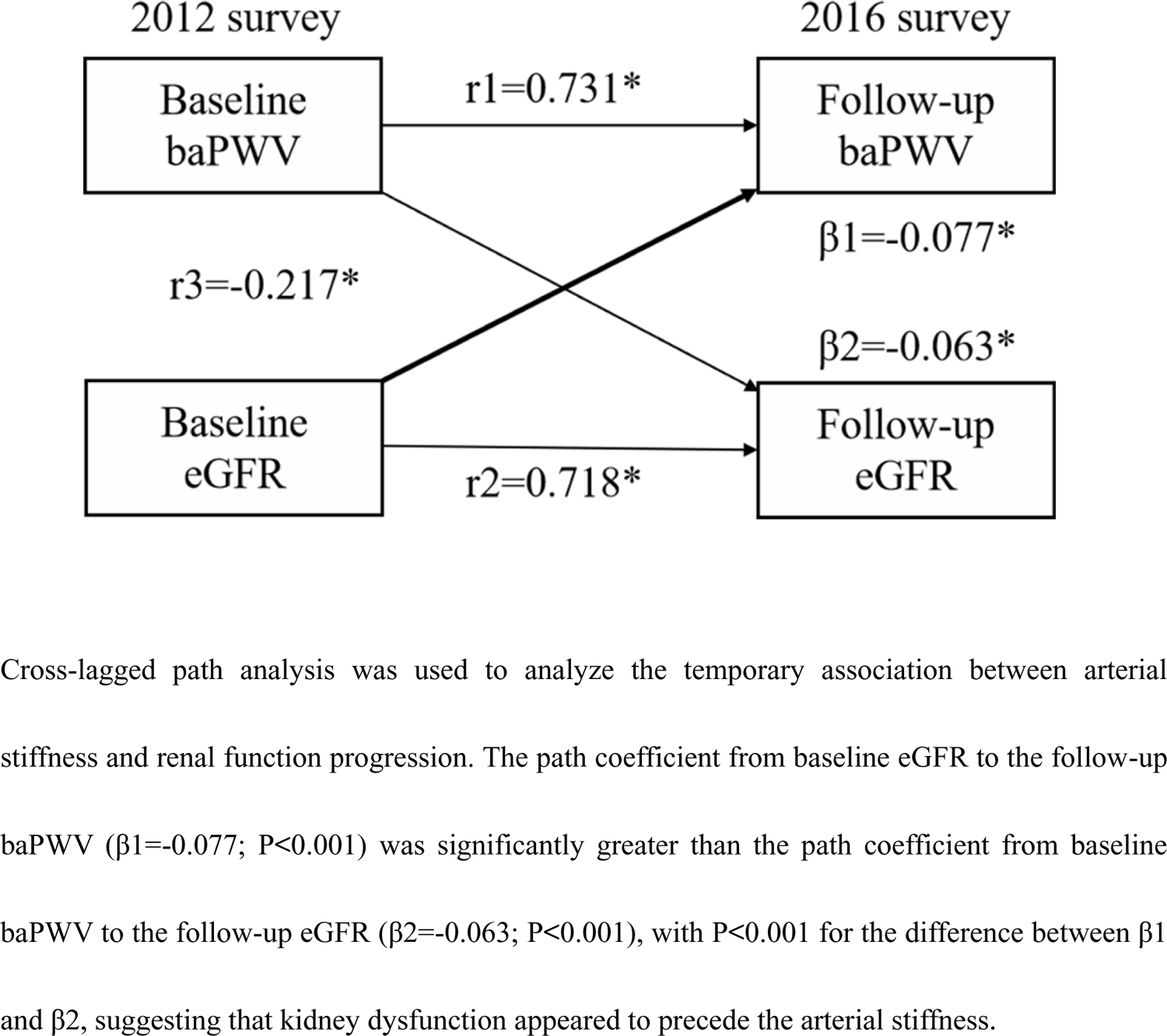
Cross-lagged analysis of brachial-ankle pulse wave velocity (baPWV) and estimated glomerular filtration rate (eGFR) (n=12334)

## DISCUSSION

This study is the first to present both cross-sectional and longitudinal associations of PWV and eGFR in a very large community-derived dataset of 67060 general adults. As a key finding, arterial stiffness was correlated to kidney function decline and its progression. Besides, there is a bidirectional connection between arterial stiffness and CKD. Arterial stiffness could increase the incidence risk for CKD, and vice versa. While through the cross-lagged analysis, it is finally showed that the kidney function decline preceded the change of arterial stiffness.

Different from our result, Varik et al. had reported a unidirectional association between arterial stiffness and CKD, that arterial stiffness amplifies renal function decline while eGFR couldn’t explain the change of PWV.^15^ Hoorn’s study and Rotterdam’s study had demonstrated eGFR was associated with arterial stiffness.^16,17^ Framingham Heart Study stated arterial stiffness had no correlates with mild-to-moderate CKD,^18^ and Magdalena Madero et al. also reported that after adjustment for systolic blood pressure the association between high aortic PWV at baseline and dynamic changes in GFR at follow-up disappeared with no statistical significance.^9^

In terms of mechanism, if the vascular lose its expansionary, the acceptance of blood flow will be reduced. Blood from the left ventricle injected into the aorta will produce a huge blood flow so that the glomerular microvascular will be damaged under the long-term perfusion of a large amount of blood flow. While the kidney can maintain normal by contraction or relaxation of the smooth muscle of the interlobular artery, and tubuloglomerular feedback or activation of the sympathetic nervous,^19^ but if the blood perfusion exceeds the self-regulation capacity of the kidney, it will be irreversibly damaged. The dysfunctional kidney also accelerates arterial stiffness through activation of the renin-angiotensin system and imbalance of calcium-phosphate metabolism. Thus, resulting in a vicious cycle.

The several strengths of our study should be noted. Our study is the first large cohort study to explore the temporal relationship between CKD and arterial stiffness by cross-lagged panel analysis. We further demonstrated the bidirectional association between CKD and arterial stiffness. Our results could provide a new direction for preventing vascular aging, as well as a new insight for health management. There are several limits to this study. First, we evaluated eGFR by serum creatinine rather than the 51Cr-EDTA technique, which is the golden standard for kidney function. Second, the observational analysis conducted in this study was mainly based on the population derived from central China, and the results need to be further verified in other regions.

## CONCLUSIONS

In summary, we show that arterial stiffness is associated with CKD. A decrease of eGFR appeared to precede the increase of baPWV.

## AUTHOR CONTRIBUTIONS

J.-J.C and F.L had full access to all of the data in the study and takes responsibility for the integrity of the data and the accuracy of the data analysis.

Concept and design: J.-J.C, F.L., and Y.L

Acquisition, analysis, or interpretation of data: J.-J.C and F.L. Drafting of the manuscript: J.-J.C and F.L

Critical revision of the manuscript for important intellectual content: Y.-M.L., J.W., R.-J.M, Z.-H.C, J.-G.W., Y.H., and Y.L

Statistical analysis: J.-J.C and F.L Obtained funding: J.-J.C, Y.H., and Y.L

Administrative, technical, or material support: Y.-M.L., J.W., R.-J.M, Z.-H.C, J.-G.W., Y.H., and Y.L

Supervision: Y.-M.L., J.W., R.-J.M, Z.-H.C, J.-G.W., Y.H., and Y.L

All authors read, critically revised and approved the final version of manuscript.

## Data Availability

Details of the study population were published in https://doi.org/10.1016/j.jacc.2019.12.039.

## ACKNOWLEDGEMENTS

None.

## Supplemental Contents

Supplemental Table 1. Baseline characteristics of the cross-sectional study

